# Geographic disparities in COVID-19 case rates are not reflected in seropositivity rates using a neighborhood survey in Chicago

**DOI:** 10.1101/2021.03.02.21252767

**Authors:** Brian Mustanski, Rana Saber, Daniel T. Ryan, Nanette Benbow, Krystal Madkins, Christina Hayford, Michael E. Newcomb, Joshua M. Schrock, Lauren A. Vaught, Nina L. Reiser, Matthew P. Velez, Ryan Hsieh, Alexis R. Demonbreun, Richard D’Aquila, Elizabeth M. McNally, Thomas W. McDade

## Abstract

To date, COVID-19 case rates are disproportionately higher in Black and Latinx communities across the U.S., leading to more hospitalizations and deaths in those communities. These differences in case rates are evident in comparisons of Chicago neighborhoods with differing race/ethnicities of their residents. Disparities could be due to neighborhoods with more adverse health outcomes associated with poverty and other social determinants of health experiencing higher prevalence of SARS-CoV-2 infection or due to greater morbidity and mortality resulting from equivalent SARS-CoV-2 infection prevalence. We surveyed five pairs of adjacent ZIP codes in Chicago with disparate COVID-19 case rates for highly specific and quantitative serological evidence of any prior infection by SARS-CoV-2 to compare with their disparate COVID-19 case rates. Dried blood spot samples were self-collected at home by internet-recruited participants in summer 2020, shortly after Chicago’s first wave of the COVID-19 pandemic. Pairs of neighboring ZIP codes with very different COVID-19 case rates had similar seropositivity rates for anti-SARS-CoV-2 receptor binding domain IgG antibodies. Overall, these findings of comparable exposure to SARS-CoV-2 across neighborhoods with very disparate COVID-19 case rates are consistent with social determinants of health, and the comorbidities related to them, driving differences in COVID-19 rates across neighborhoods.

---

The US has led the world in numbers of SARS-CoV-2 infections (1) with Chicago as one of the epicenters with 241,655 cases of COVID-19 resulting in 4,909 deaths by February 20, 2021 (2). From early in the pandemic, stark racial/ethnic disparities were evident in COVID-19 deaths. Subsequent analyses of public health surveillance data further explicated the nature of COVID-19 racial disparities, with many studies showing that communities with more Black or Latinx residents had higher case and mortality rates (3). Studies examining the drivers of these racial/ethnic disparities have found relatively consistent associations with residential segregation of economic disadvantage, proportion of essential workers, and crowded living conditions (4-6). There have been inconsistent results across studies regarding whether comorbidities mediate the observed racial/ethnic disparities (3, 7, 8).

The analyses demonstrating such disparities mostly relied on molecular testing to diagnose acute, symptomatic COVID-19 illness by detecting SARS-CoV-2 RNA in nasal and nasopharyngeal swabs (9). However, from the beginning of the pandemic until the present, acute viral diagnostic testing has not always been readily accessible to all. This could limit accurate estimation of the COVID-19 case rate and epidemiological patterns of SARS-CoV-2 spread by not identifying minimally symptomatic and asymptomatic, but potentially infectious, cases (10-13). Many infections are asymptomatic or minimally symptomatic, although still capable of transmitting SARS-CoV-2 to others (10). A complementary epidemiological approach is serological testing to detect antibodies against the virus (14, 15). Virus protein-specific antibodies emerge within weeks after infection, and subsequent IgG specific antibodies remain detectable for 4 or more months after infection, depending on assay sensitivity and specificity. This is much longer than the several weeks that airway virus RNA remains detectable (16-19) after recovery from symptoms (20). Thus, serology has portential to identify infections long after their initiation, independent of clinical diagnosis and symptom experience, potentially providing a fuller epidemiologic assessment of the pandemic. Assessment of all SARS-CoV-2 exposures by antibody detection can also test the hypothesis that risk of the viral infection itself varies by race/ethnicity, rather than only a subsequent COVID-19 illness being associated with race/ethnicity.

Fewer seroprevalence studies of racial and geographic disparities have been reported to date, relative to comparisons of virus RNA-based COVID-19 case surveillance. These studies suggest there are racial/ethnic disparities in SARS-CoV-2 infection, although they have some limitations and inconsistent results. One early study of fingerstick blood specimens collected at grocery stores in New York state observed significantly higher prevalence of IgG antibodies to the SARS-CoV-2 nucleocapsid protein, assessed as detectable versus not detectable antibody, among Latinx, Black, and Asian adults relative to white adults (21). Another study in Baton Rouge using the qualitative Abbott SARS-CoV-2 anti-nucleocapsid IgG nucleoprotein enzyme-linked immunosorbent assays (ELISA) assay (ARCHITECT™ platform) was not indubitably confirmatory, as it found much smaller differences. Weighted seroprevalence was 9.8% among participants who were Black, 7.1% multiracial, 5.5% Asian, 4.5% White, and 5.3% Hispanic; confidence intervals were non-overlapping except for Asian and Hispanic (22). Both reports had the limitation that the qualitative anti-nucleocapsid antibody assays used have been reported to not be as sensitive and specific as quantitative ELISA detection of anti-SARS-CoV-2 spike glycoprotein receptor binding domain (RBD) IgG. Previous reports suggest that an anti-RBD IgG assay exhibits superior performance compared to an anti-nucleocapsid IgG assay (19, 23-26). These two earlier assessments of racial/ethnic disparities in seroprevalence also relied on in-person collection of samples at a clinical care or research site, which limits reach and coverage during varying pandemic mitigations such as stay at home orders similarly to acute virus diagnostic testing.

In Chicago, like many cities, there are large differences in documented rates of COVID-19 cases and fatalities across neighborhoods that vary in their racial/ethnic and socioeconomic composition (2). However, the lack of comprehensive seroprevalence assessments reported to date has not yet enabled analysis of whether SARS-CoV-2 infections are similarly disparate. The current study tested the hypothesis that geographic disparities in COVID-19 case rates will be mirrored in seroprevalence rates, using a more sensitive, specific, and quantitative antibody assay than the earlier reports (21, 22). Moreover, the antibody assay used here is not dependent upon access to in-person testing at care or research sites. To achieve this, we utilized dried blood spots (DBS) collected in the home setting using a simple finger prick method. This followed web-and mass media-facilitated participant recruitment and online screening/consenting. After blood collection, participants’ DBS cards were returned to the laboratory for testing in pre-paid mailers. We used a previously described the DBS-based quantitative ELISA for IgG to the receptor binding domain of the SARS-CoV-2 protein (27). Our study design offered a strong opportunity to confirm the hypothesis that geographic disparities would be similar in molecular diagnostic and serosurveillance assays across 5 pairs of Chicago ZIP codes that both bordered one another and had very different COVID-19 case rates. Results instead suggest there are differences in COVID-19 illness between these neighborhoods, but not SARS-CoV-2 infection.

## METHODS

All research activities were implemented under conditions of informed consent with protocols approved by the institutional review board of the university where the authors are primarily affiliated. Study data were collected and managed using REDCap electronic data capture tools (28).

To select ZIP code pairs, COVID-19 cumulative case rates by ZIP code were obtained from Chicago Department of Public Health’s COVID-19 website (2). Additional health, socio-economic, and demographic ZIP code level data were selected from US Census Bureau American Community Survey (29) and the Chicago Health Atlas (listed in Table 2) (30). ZIP codes were ranked based on the CDPH COVID-19 cumulative case rate for the week of 4/26/20-5/2/20. Low case rate ZIP codes (category 1 or 2) that were adjacent to high case rate ZIP codes (category 4 or 5) were selected for review. In cases were multiple touching ZIPs met the criteria, a ZIP was selected based on additional criteria listed in Table 2. Five ZIP code pairs were chosen based on geographic location, highest case rates, and these additional socio-economic, health, and demographic characteristics.

**Table 1:** Demographic characteristics of sample overall and by antibody serostatus, Chicago IL, 2020 (n=1,667)

|  | Full Sample<br>n (Col %) | Seropositive<br>n (Row %, 95% CI) | Seronegative<br>n (Row %, 95% CI) |
| --- | --- | --- | --- |
| Age |  |  |  |
| 18-29 | 300 (18.0) | 59 (19.7, 15.2-24.2) | 241 (80.3, 65.8-84.8) |
| 30-39 | 542 (32.5) | 95 (17.5, 14.3-20.7) | 447 (82.5, 79.3-85.7) |
| 40-49 | 366 (22.0) | 83 (22.7, 18.4-27.0) | 283 (77.3, 73.0-81.6) |
| 50-59 | 252 (15.1) | 45 (17.9, 13.1-22.6) | 207 (82.1, 77.4-86.9) |
| 60-69 | 146 (8.8) | 20 (13.7, 8.1-19.3) | 126 (83.6, 80.7-91.9) |
| 70+ | 61 (3.7) | 10 (16.4, 7.1-25.7) | 51 (83.6, 74.3-92.9) |
| Gender |  |  |  |
| Male | 707 (42.4) | 125 (17.7, 14.9-20.5) | 582 (82.3, 79.5-85.1) |
| Female | 954 (57.2) | 187 (19.6, 17.1-22.1) | 767 (80.4, 77.9-82.9) |
| Transgender | 6 (0.4) | -- | -- |
| Race/Ethnicity |  |  |  |
| Asian, non-Latinx | 184 (11.0) | 29 (15.8, 10.5-21.0) | 155 (84.2, 79.0-89.5) |
| Black, non-Latinx | 142 (8.5) | 30 (21.1, 14.4-27.8) | 112 (78.9, 72.2-85.6) |
| Latinx | 386 (23.2) | 100 (25.9, 21.5-30.3) | 286 (74.1, 69.7-78.5) |
| Other, non-Latinx | 50 (3.0) | 12 (24.0, 12.2-35.8) | 38 (76.0, 64.2-87.8) |
| White, non-Latinx | 905 (54.3) | 141 (15.6, 13.2-17.9) | 764 (84.4, 82.1-86.8) |
| Prior COVID-19 testing<sup>2</sup> |  |  |  |
| Never tested | 1188 (71.4) | 204 (17.2, 15.0-19.3) | 984 (82.8, 80.7-85.0) |
| Tested negative | 436 (26.2) | 70 (16.1, 12.6-19.5) | 366 (83.9, 80.5-87.4) |
| Tested positive | 41 (2.5) | 37 (90.2, 81.2-99.3) | 4 (9.8, 0.7-18.8) |
| Worked outside home since March<sup>3</sup> |  |  |  |
| Yes, healthcare | 202 (12.1) | 39 (19.3, 13.9-24.8) | 163 (80.7, 75.2-86.1) |
| Yes, other | 427 (25.7) | 78 (18.3, 14.6-21.9) | 349 (81.7, 78.1-85.4) |
| No, working from home | 669 (40.2) | 112 (16.7, 13.9-19.6) | 557 (83.3, 80.4-86.1) |
| No, not employed | 365 (21.9) | 83 (22.7, 18.4-27.0) | 282 (77.3, 73.0-81.6) |

<sup>1</sup>Values suppressed to prevent identifiability due to small cell sizes.
<sup>2</sup>Missing data for two subjects.
<sup>3</sup>Missing data for four subjects.

**Table 2:**
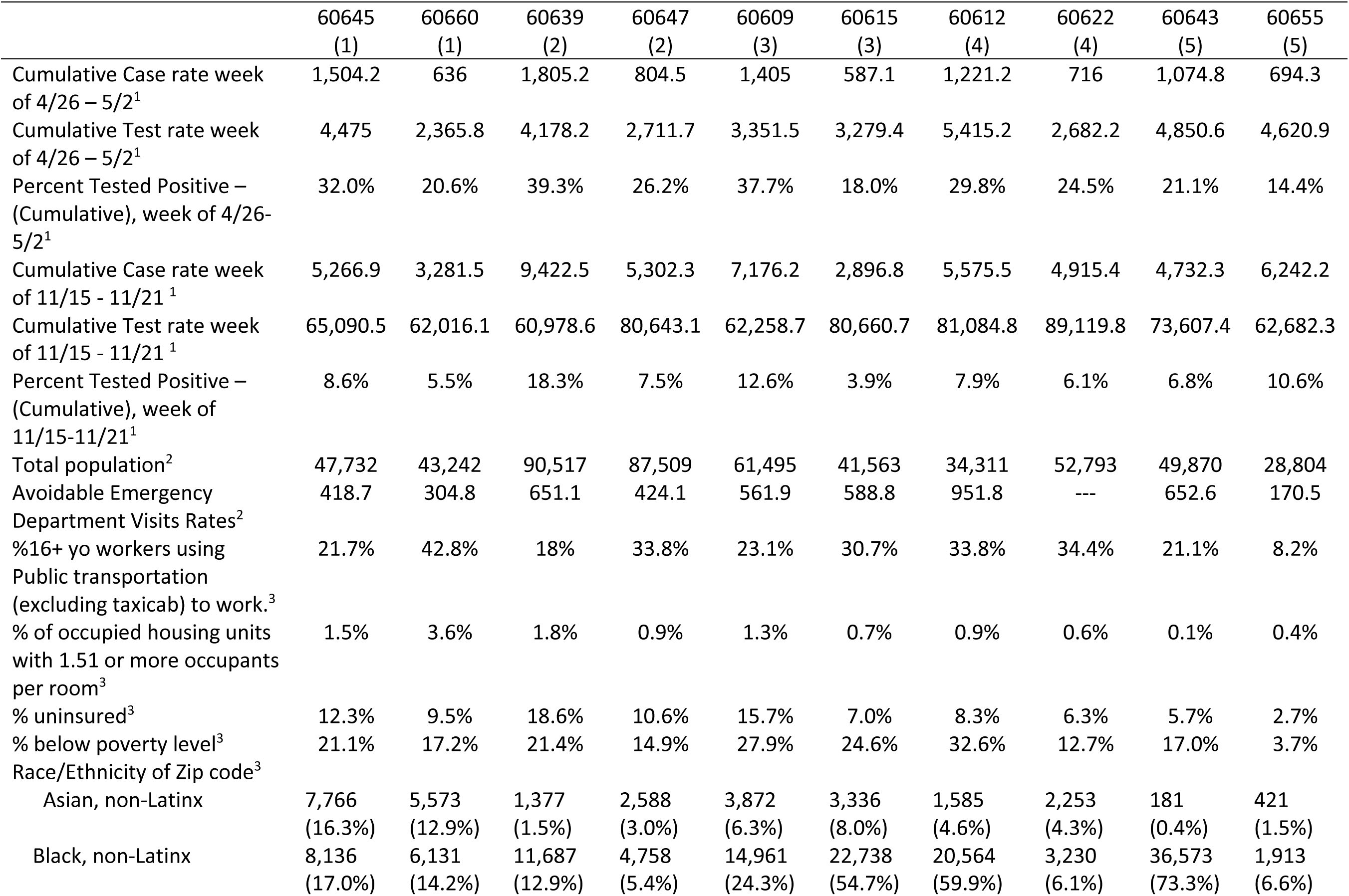
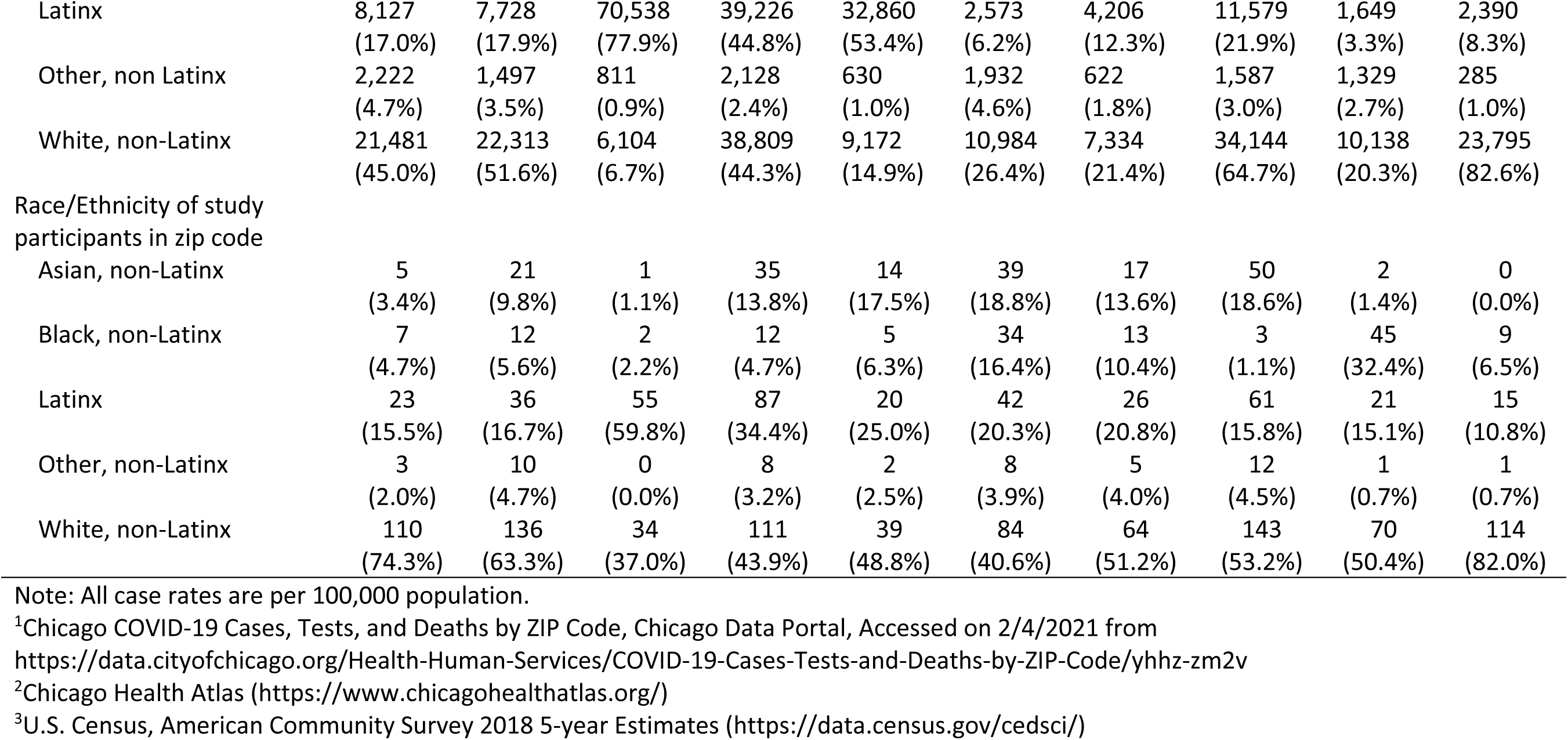
Characteristics of zip code pairs, Chicago, IL

### Procedures

To achieve a diverse representation of residents of these ZIP codes, participants were recruited through two mechanisms. First, community-based participants (n = 1,509) were recruited from ten ZIP codes in Chicago through social media advertising, outreach to local businesses and community leaders, and news articles about the study. Participants were screened for eligibility, which included ZIP code residence and age of 18 years or over. To assure racial and gender diversity within the sample, enrollment of women and white participants (groups that disproportionately completed the screener) was adaptively matched to enrollment of men and racial-minority participants within each ZIP code. Second, staff, students and faculty from the Northwestern University’s Feinberg School of Medicine (FSM) in Chicago, IL were sent an email describing the study with a link to the website (n = 158 who lived in one of the 10 study ZIP codes). Eligible participants were invited to complete a questionnaire regarding health status, including COVID-19 symptoms. Community participants received materials for DBS collection through the United States Postal Service (USPS) and returned their test kits using prepaid USPS envelopes provided to them by the study team. Those affiliated with FSM were given a specific time to collect DBS kits in person and were instructed to return their completed kits to the same location. The research team developed a video that explained to participants all of the steps of DBS collection and return (31). Sample collection occurred between June 24 and November 23, 2020.

We measured antibodies to RBD because previous studies have reported better sensitivity and specificity for SARS-CoV-2 infection detection than seen with anti-nucleocapsid antibodies (19, 23-26). Anti-RBD antibodies have infrequent and low-level cross-reactivity to other seasonal human coronaviruses (32, 33), an advantage relative to detection of antibodies to other SARS-CoV-2 proteins. The ELISA protocol we used has been validated for DBS and previously described (27, 34). Samples were run in duplicate and reported as the average. Results were normalized to the CR3022 antibody with known affinity (35). A value >0.39µg/ml CR3022 was considered positive. Statistical analysis

Unadjusted and adjusted binomial logistic regression was performed to compare seroprevalence within each ZIP code pair. Age, race/ethnicity, gender and prior COVID-19 testing result were used as covariates when reporting adjusted odds ratios. Geographical information system (GIS) analyses were conducted using ArcGIS Pro 2.6.2. Only participants who were geocoded using the StreetAddress or AddressPoint locator and were geocoded to the ten ZIP codes chosen for analysis were included in the geospatial analysis (n = 1,666). Data sources for GIS data are indicated in Table 2.

## RESULTS

Table 1 reports sample characteristics. A slight majority of the sample was non-Latinx white (54.3%), most had never tested for COVID-19 (71.4%), and the sample was heterogenous in terms of employment-based SARS-CoV-2 exposure risk. Overall, the seroprevalence in the sample was 18.7%. As indicated by non-overlapping confidence intervals in estimates in Table 1, seroprevalence was significantly higher in Latinx (25.9%) than White non-Latinx (15.6%), and in those who had previously tested COVID-19 positive (90.2%) than those who had never tested (17.2%) or tested negative (16.1%).

Figure 1 shows a map with the distribution of seropositive and seronegative negative participants within the 10 ZIP codes. Finer grain maps of participants’ residential locations with geographic features overlayed (e.g., highways, train yards, waterways, etc.; not shown to protect participant confidentiality) suggested an even distribution of participants within residential spaces in each ZIP code, with the exception of 60643 in which there were few cases in residential areas east of the interstate highway. This even distribution suggests a lack of clustering of participants along paired ZIP code boundaries in a way that would have confounded our analyses of similarities between case rates and seroprevalence. Multiple approaches to geospatial hotspot analyses did not demonstrate significant cluster of seroprevalence within ZIP codes.

**Figure 1:**
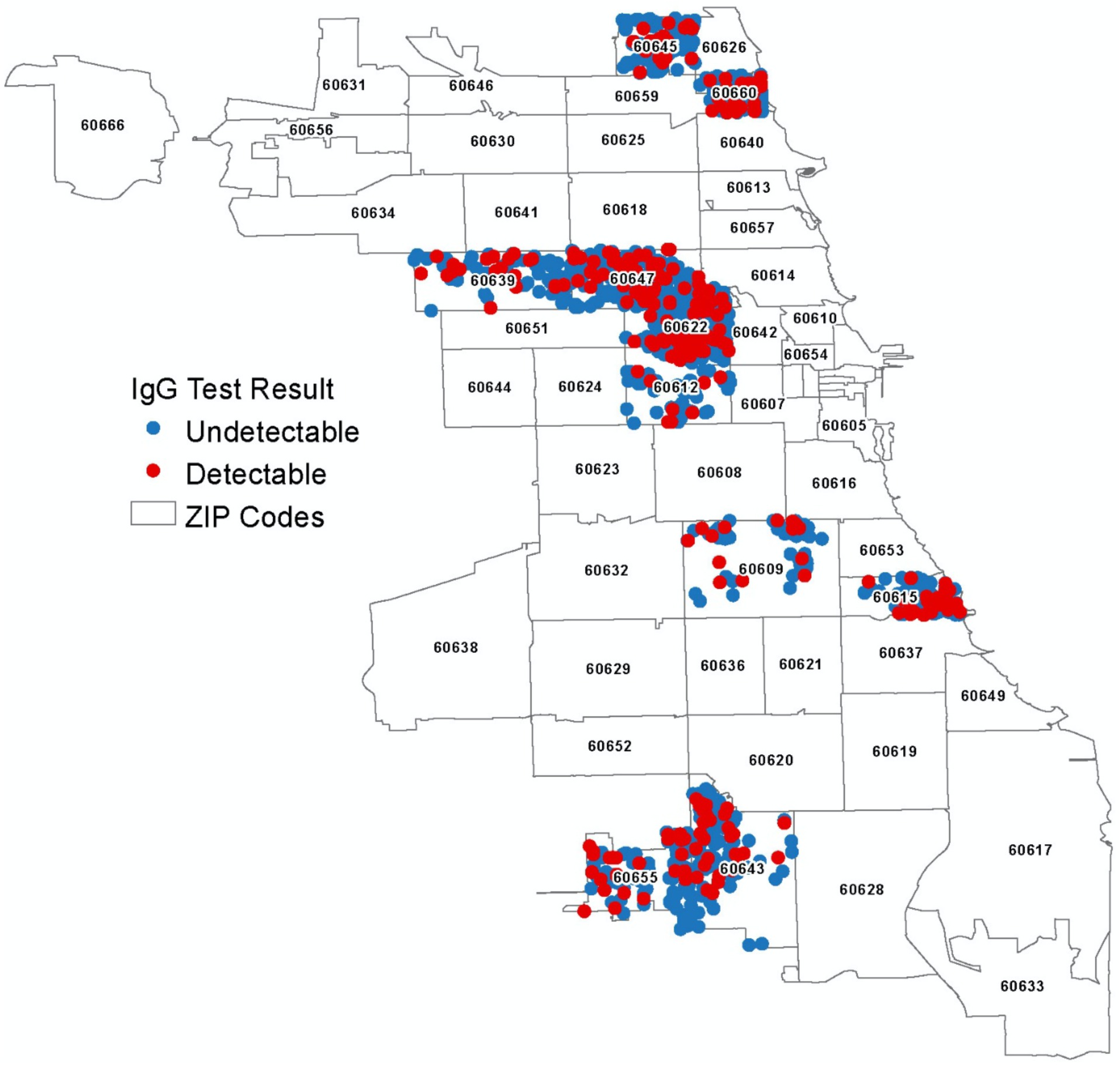
GIS plot of SARS-Cov-2 cases within 10 zip codes included in the sampling frame, Chicago, IL 2020. Source: ESRI, TomTom North America, Inc, United States Postal Service

Table 2 presents characteristics of the 10 ZIP codes included in the study. In each pair, the ZIP code with the higher case rate is in the column to the left of the ZIP code with the lower case rate. Cumulative case rates in all ZIP codes increased from the time when ZIP codes were first selected to the end of data collection, and most ZIP code pairs were relatively stable in terms of the ratio of reported cases across pairs, with the exception of pair 5 (60643 and 60655). In this pair, 60655 had a much larger increase in the case rate than any other ZIP code. Rates of COVID-19 testing by ZIP code were not available at the time of ZIP code selection, but were subsequently released. Across pairs, cumulative test numbers in April/May were higher (pairs 1, 2, 4) in ZIP codes with more cases or similar across pairs (pairs 3, 5). By November, number of cumulative tests became more similar across pairs. Zip code pairs varied considerably in certain observed characteristics, such as population size, markers of healthcare accessibility (i.e., rates of avoidable emergency department visits), transportation-based exposure risk (i.e., use of public transportation to work), poverty, and race/ethnicity. Race/ethnicity of study participants also varied by ZIP code, but non-white participants were underrepresented in each. Better representation was achieved with Latinx participants.

Figure 2 plots SARS-CoV-2 seroprevalence vs. cumulative COVID-19 case rates for each ZIP code pair. Table 3 shows odds ratios of seroprevalence within each of the 5 ZIP code pairs, none of which were significant; even trends across pairs did not support the hypothesis that adjacent ZIP codes with higher case rates trended towards a higher seroprevalence based on antibody testing. Adjustments with individual-level covariates that differ across ZIP codes (e.g., race/ethnicity) did alter some point estimates but did not change the pattern of significance of results. Analyses weighting data to census characteristics (not shown) also did not alter the pattern or significance of findings.

**Figure 2.**
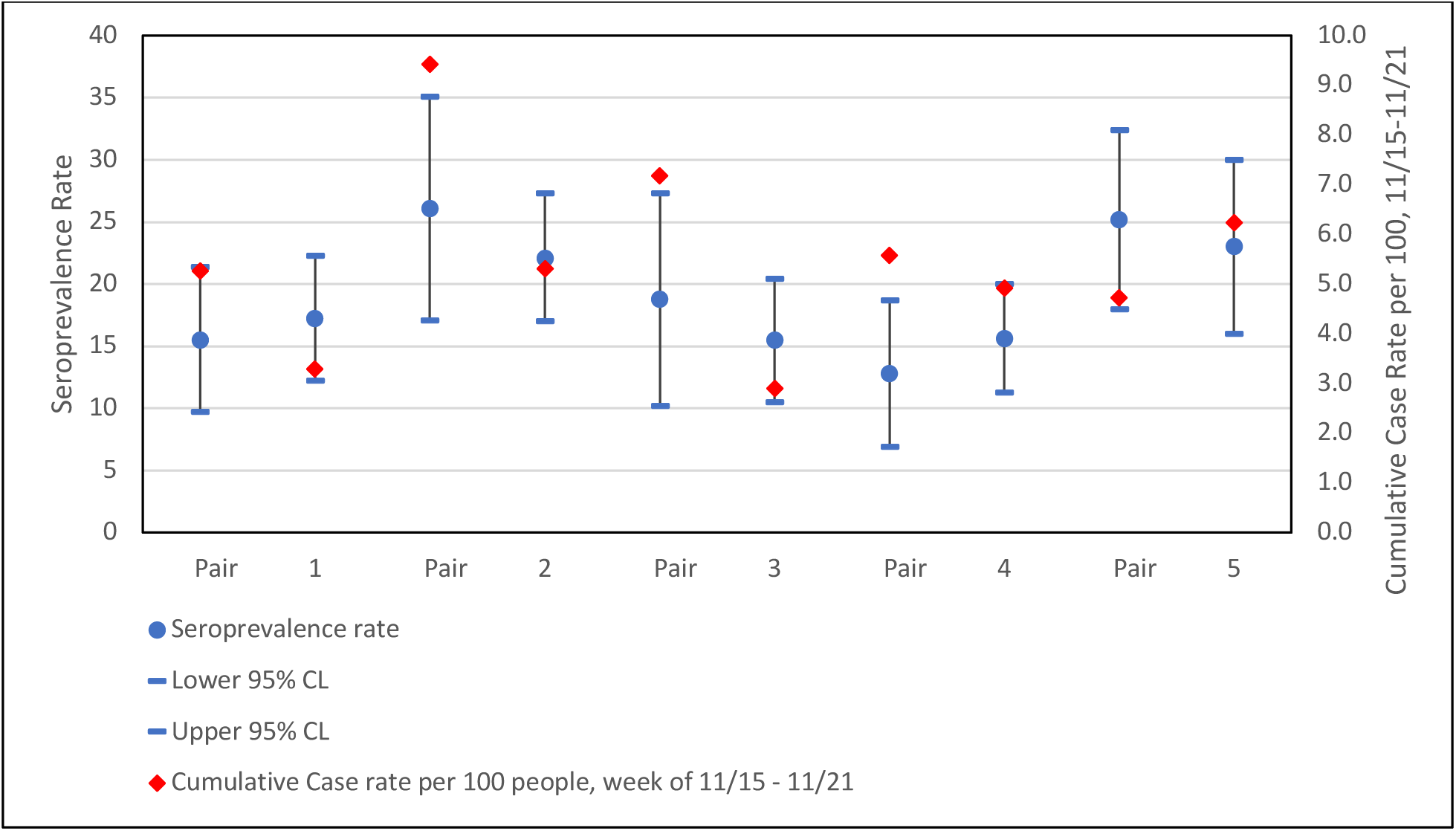
SARS-Cov-2 Seroprevalence rates vs. cumulative COVID-19 case rate by ZIP code pair. Adjacent ZIP codes with substantial differences in cumulative case rates were selected and surveyed for seroprevalence. We did not detect statistically significant differences in seroprevalence between the high case rate ZIP code and the low case rate ZIP code for any of the pairs surveyed. This suggests that the prevalence of SARS-Cov-2 exposure does not account for neighborhood differences in COVID-19 case rates.

**Table 3:** Unadjusted and adjusted comparisons of seroprevalence within zip code pairs, n = 1,667, Chicago, IL 2020

|  | OR (95% CI) | aOR (95% CI) |
| --- | --- | --- |
| Pair 1 (60645 60660) | 0.89 (0.50, 1.56) | 0.96 (0.53, 1.71) |
| Pair 2 (60639 60647) | 1.24 (0.71, 2.16) | 1.07 (0.60, 1.91) |
| Pair 3 (60609 60615) | 1.26 (0.64, 2.48) | 1.21 (0.60, 2.41) |
| Pair 4 (60612 60622) | 0.79 (0.43, 1.47) | 0.73 (0.39, 1.37) |
| Pair 5 (60643 60655) | 1.13 (0.65, 1.95) | 0.91 (0.51, 1.61) |

Note: CI = confidence interval; OR = Odds Ratio; aOR = Adjusted Odds Ratio Logistic regression analyses included the following covariates to produce adjusted ORs: race/ethnicity, age, gender, and prior COVID-19 positive test. All effects non-significant with $p > .05$ .

## DISCUSSION

In this study we sought to test the hypothesis that geographic differences in epidemiology of COVID-19 based on case reports would be confirmed in a seroprevalence study of IgG antibodies to the SARS-CoV-2 spike protein. Our comparison of 5 adjacent ZIP codes with substantially different COVID-19 case rates at the time of selection did not confirm this hypothesis. In unadjusted, adjusted, and weighted analyses, there were no significant differences in seroprevalence between ZIP codes in the same pair. This suggests little association between case rates by ZIP code and infection rates as estimated by serosurveillance.

There are several possible explanations for why seroprevalence may not track case rates. First, it is possible that our seroprevalence study provided a better representation of SARS-CoV-2 epidemiology than case rates. We used a highly sensitive assay (27), and therefore likely detected positive individuals who were asymptomatic and never sought testing. At times, diagnostic testing in Chicago was unavailable or extremely restricted, and therefore even symptomatic individuals were not tested; these untested individuals would have been identified as having had COVID-19 with this antibody screen. Further, participants in the current study could be screened for antibodies by providing a sample they collect at home, and even when diagnostic testing was available, some individuals feared leaving their home to go to testing spaces. Along these same lines, serological antibody testing is a marker of past infections, whereas PCR diagnostic testing only detects active infections. If serology is indeed a more accurate representation of SARS-CoV-2 epidemiology, it would suggest that exposure to SARS-CoV-2 may have been more widespread than is indicated by case rates. This conclusion is also supported by a recent study comparing seroprevalence to case rates in 10 sites in the U.S. (36).

Second, it is possible that, due to the sensitivity of the assay used in our study, we detected individuals who had lower doses of exposure to SARS-CoV-2 virus at the time of exposure, which may be associated with an asymptomatic or less intense clinical presentation (37, 38). If this is true, then observed variation in case rates across ZIP codes may actually represent differences in severity of illness. Individuals with a larger exposure doses may have an increased likelihood of experiencing symptoms and subsequently receiving diagnostic testing. Thus, individuals across these ZIP code pairs may not differ in their rates of actual exposure, but instead in their dose of exposure. If this is the case, it suggests that the use of serosurveillance studies to inform public health action may need to consider the disconnect between detection of any exposure and clinical risk. Quantitative antibody assays linking levels of antibody response to clinical presentation may be particularly useful for such research. At the same time, larger exposure doses would likely be tied to a close proximity between an infected and uninfected individual, such as those sharing a room in the same home. While 10 ZIP codes is too few for a formal statistical test, in our study a widely used metric of residential crowding (% of housing with 1.51+ occupants per room) showed little-to-no association with ZIP code seroprevalence. Of course, ZIP code level living density is not synonymous with density assessed at the level of a single home, so future research should assess number of people in home and in a shared bedroom.

A third possibility is that case rates are closely tied to symptoms, and symptoms are tied to older age and underlying comorbidities (39), so even in conditions of constant levels of exposure/infection/transmission across neighborhoods, those with more social determinants of chronic diseases would tend to have higher case rates— assuming equal access to testing. When study ZIP codes were selected in April/May, testing as a proportion of the population was low across all ZIP codes (mean = 8.3% range 3.1-16.0%), but increased until at study completion most ZIP codes had more cumulative tests than residents. While the current sample size of 10 ZIP codes is too small to compute a correlation, the pattern of results does not suggest that diagnostic testing rates are positively associated with seropositivity. Census markers of poor healthcare access also did not show a strong positive trend with seropositivity. Admittedly, these are crude markers of presence of chronic diseases so future studies should examine additional markers. Overall, this pattern is consistent with more uniform spread of infections, including those that are asymptomatic, across Chicago ZIP codes than would be suggested by case surveillance data.

Our study also examined the relationship between individual characteristics and seropositivity. Differences by age, gender groups, and employment status stratified by possible exposure risk were not significant. As an epidemiological confirmation of the validity of the antibody assay used in this study, participants who self-reported a prior positive COVID test were substantially more likely to be seropositive than those who did not (90.2% versus 16.1-17.2%). Most racial minority groups trended toward higher seropositivity, but the only significant difference was between Latinx and White, non-Latinx participants (25.9% versus 15.6%). These findings are consistent with case surveillance in Chicago, which indicated that at the time of the launch of data collection, case rates were surging in the Latinx community. As of February 5, 2021, the infection case rate in the Latinx population was higher than non-Latinx White and Black combined (10,797.4, 4,851.2, and 5,227.8, respectively)(2). Test positivity rates showed a similar pattern. Cumulative deaths since the beginning of the pandemic have occurred substantially more among Black Chicagoans; in fact, this alarming pattern is what inspired the focus of this study seeking to understand disparities in SARS-CoV-2 spread. Fortunately, these inequities in deaths from COVID-19 have shrunk over time as death rates overall have declined in the context of more effective prevention and treatment. This pattern is consistent with some partial role of pre-existing comorbidities in explaining racial disparities in COVID-19 disease and death; the magnitude or existence of such an effect is still contested in the existing literature and will require further research (3, 7, 8). Such an effect would support repeating again the call to urgently address social determinants of health that produce inequities in chronic diseases that will exacerbate medical disparities when pandemic occur (40, 41).

Findings must be considered in the context of study limitations. This was not a probability sample, but instead obtained through social and news media, community outreach, and employees of a large medical school. Our goal was not to provide population estimates of seroprevalence, rather we sought to look at relative rates within pairs of carefully selected adjacent ZIP codes with differing case rates. Nevertheless, our estimation of seroprevalence within ZIP codes may be biased by our sampling approach. For example, despite attempts to target recruitment from Black communities, the proportion of Black participants in the study underrepresented the demographics of Chicago. We sought to partially correct for this with sensitivity analyses that included demographic covariates and weighting. Further, individuals who opted to volunteer to participate when reading about the study through widespread coverage in local news, social media ads, or community outreach may not represent the entire community. Second, participants were required to self-collect a DBS sample. Through well-produced videos and collection materials we hoped to minimize concerns, but selection biases may still have operated.

## Conclusion

Our findings indicate that exposure to SARS-CoV-2 may be more consistent across neighborhoods within Chicago than was previously thought based on reported COVID-19 case rates. This suggests that factors other than differential seroprevalence may play a role in driving disparities in COVID-19 outcomes. One possibility is that pre-existing chronic conditions are associated with greater risks of symptomatic infection, leading to higher rates of symptomatic illness and case detection in groups with higher rates pre-existing chronic conditions. Another possibility is that the average dose of exposure is higher in some neighborhoods compared to others, leading higher rates of symptomatic illness and case detection in areas where the intensity of exposure is greater. Differences in viral dose may occur for a variety of reasons including differences in adherence to preventive behaviors, work environments, or living situations. Our results highlight the importance of investigating other factors besides differential exposure as potential drivers of inequity in COVID-19 outcomes.

## Data Availability

Data are available upon request from the corresponding author subject to an approved data sharing agreement to protect participant confidentiality.

## Notes

### Competing Interest Statement

Dr. McDade reports that he has a financial interest in EnMed Microanalytics, a company that conducts lab tests using DBS samples.
Dr. D'Aquila reports personal fees from Abbvie, outside the submitted work.
Dr. McNally reports personal fees from Amgen, personal fees from AstraZeneca, personal fees from Cytokinetics, personal fees from Pfizer, personal fees from Tenaya Therapeutics, personal fees from 4D Molecular Therapeutics, outside the submitted work.

### Funding Statement

This material is based upon work supported by the National Science Foundation under Grant No. 2035114. Additional support was provided by the Northwestern University Office of Research, the Northwestern University Clinical and Translational Sciences Institute (NIH UL1TR001422), and a generous gift from Dr. Andrew Senyei and Noni Senyei. The funding sources had no role in the study design, data collection, analysis, interpretation, or writing of the report.

### Author Declarations

All samples were de-identified and all research activities were implemented under protocols approved by the institutional review board at Northwestern University (#STU00212457 and #STU00212472).

